# Does active travel behaviour moderate the association between the built environment and depression?

**DOI:** 10.64898/2026.10.08.26364859

**Authors:** Faye Sanders, Vilte Baltramonaityte, Lorraine Whitmarsh, Esther Walton

## Abstract

**Background:** Built environment characteristics have been associated with depressive symptoms, whilst active travel behaviour is linked to improved mental health. However, it remains unclear whether active travel buffers associations between increased exposure to the built environment and worse mental health.

**Methods:** In this pre-registered study, participants were 10,310 mothers (mean age = 28.11, SD = 4.83) participating in the Avon Longitudinal Study of Parents and Children (ALSPAC). A latent built environment factor was measured at study enrolment, active travel behaviour at a 2-year follow-up, and depressive symptoms at a 4-year follow-up. Moderation was examined using adjusted linear regression models including an interaction term between the built environment factor and active travel behaviour.

**Results:** The built environment factor was positively associated with depressive symptoms (β = 0.023, *p* = .012). Active travel behaviour was not associated with depressive symptoms (β = −0.002, *p* = .854) and did not moderate the association between the built environment factor and depressive symptoms (β = −0.002, *p* = .865), after adjusting for covariates, including age, familial depressive history, socioeconomic risk factors, physical activity, baseline depressive symptoms and smoking.

**Conclusion:** Contrary to expectations, active travel behaviour does not modify associations between the built environment and depressive symptoms. These findings provide no evidence that active travel mitigates mental health risks associated with the built environment in this population.

**What is already known on this topic:**

- Some features of the built environment have been associated with poor mental health, and active travel has been linked to better psychological wellbeing. However, it is unclear whether active travel can reduce the mental health risks associated with more urbanised environments.

**What this study adds:**

- We tested active travel as a potential modifier of the longitudinal association between the built environment and depressive symptoms.
- We found no evidence that active travel buffered this association in a sample of mothers.

**How this study might affect research, practice or policy:**

- Engagement in active travel alone may be insufficient to offset mental health risks associated with the built environment, and future research should test whether these relationships differ across populations, travel purposes and levels of active travel.

## Introduction

There are multiple factors within the built environment that associate with poor mental health. For example, previous research examining single features of the built environment with mental health, such as population density, have found associations between increased rates of psychiatric disorders and increased population density [1]. In addition, some features of the built environment, including accessible and high-quality green space, have been found to be protective of both depressive and anxiety symptoms [2].

Research modelling the interconnectedness of built environment features has shown more complex relationships with depressive symptoms. For example, whilst feature-specific associations with depressive symptoms in network analyses have been identified as generally weak, an overall built environment factor, including features such as population density, building density, green space and walkability, appeared to associate more strongly and consistently with increased depressive symptoms in previous work [3]. This suggests that a built environment factor may explain depressive symptoms better when features are considered collectively than in isolation. Moreover, whilst the association between a built environment factor and depressive symptoms was strongest cross-sectionally, this association was also present over a 4-year follow-up period, highlighting the importance of longitudinal research when assessing relationships between the built environment and mental health. Whilst this previous research has advanced our understanding of associations between built environment and depression, our understanding of factors that may modify this relationship are still poorly understood.

One factor which may provide promising protective effects on associations between the built environment and depression is active travel behaviour. Active travel behaviour refers to the ways in which individuals use physical energy expenditure on a journey, and includes activities such as walking, cycling and skateboarding and excludes transport modes like driving [4]. Previous research often focused on individual characteristics such as pro-environmental attitudes or personality traits to predict active travel behaviour [5,6], and found that interventions targeting these factors have shown limited effectiveness and longevity in behaviour change [7]. This suggests predictors of active travel behaviour are likely more complex than individual factors alone. More recently, studies have indicated that the built environment may associate with individuals’ travel behaviour [8,9]. For example, the transport environment, including supportive infrastructure, associates with active travel behaviour [10,11]. In fact, those living in an urban area with greater walkability have been reported to be over twice as likely to walk or cycle than those living in a rural area, explaining greater variance in active travel behaviour than sex, age, education, income and health status [11]. Similarly, evidence from the UK Household Longitudinal Survey found that urban residents were 64% more likely to use active travel than rural residents [12]. Recent studies have also addressed the concern of residential self-selection (the idea that individuals who prefer walking or cycling may deliberately choose to live in more walkable neighbourhoods), by statistically controlling for factors such as socio-demographic characteristics (e.g., age, income, education) and personal attitudes that might influence relocation decisions. Even after accounting for these potential biases, the studies still found a strong link between walkable urban environments and increased rates of active travel, such as walking or cycling [8,13], evidencing further the importance of the built environment for differences in active travel between urban and rural settings.

Active travel behaviour may play a substantial role in modifying mental health risk associated with the built environment. Active travel interventions have shown significant benefits for mental wellbeing, including mental health functioning and perceived personal wellbeing [14]. Longitudinal evidence further suggests the association between active travel behaviour and mental health is not simply driven by reverse causation [15]. Unlike bidirectional and reverse associations between active travel and physical health, whereby those with better physical health are more likely to engage in active travel behaviours, cross-lagged panel analyses suggest relationships between active travel behaviour and mental health, including affective components of depression, are less likely to be driven by reverse relationships [15]. In other words, these analyses suggest that mental health is more likely an outcome of active travel than a predictor.

Whilst it appears that there are indeed associations between the built environment, active travel behaviour and mental health, it remains unclear as to whether associations between the built environment and depression vary according to levels of active travel behaviour. Urban living is generally associated with increased risk for depression [1], while active travel is linked to better mental health [15]. Understanding whether active travel behaviour could act as a protective factor to mitigate urban-related mental health risks is critical. Therefore, using longitudinal data from ALSPAC (Avon Longitudinal Study of Parents and Children) this study aims to investigate whether associations between the built environment and depression can be moderated by active travel behaviour.

In our pre-registration, we hypothesised that active travel behaviour would moderate the association between the built environment and depressive symptoms, such that the risk association between built environment and depressive symptoms would be weaker among individuals with higher levels of active travel.

## Methods

### Participants

Participants were mothers from the Avon Longitudinal Study of Parents and Children (ALSPAC) cohort [16–18]. Pregnant women resident in Avon, UK with expected dates of delivery 1st April 1991 to 31st December 1992 were invited to take part in the study and the initial number of pregnancies enrolled was 14,541. We selected mothers who had at least 50% data available for built environment features (including building density (300m buffer), connectivity density (300m buffer), facility density (300m buffer), facility richness (300m buffer), bus lines length (300m buffer), bus stops (500m buffer), total traffic load (100m buffer), distance to nearest road, walkability (300m buffer), green space (500m buffer) and population density) and depressive symptoms at study enrolment, leaving up to 10,310 mothers available for analysis. We intended to also include bus stops and traffic load as built environment features, but this was not possible due to missing data (99.5% and 91.5% respectively). The study website (https://www.bristol.ac.uk/alspac/researchers/our-data/) contains details of all data available through a fully searchable data dictionary and variable search tool.

Ethical approval for the study was obtained from the ALSPAC Ethics and Law Committee and the Local Research Ethics Committees (IDs: E1808, 49/89 and 90/8) and Department of Psychology at the University of Bath (6992-9812). Informed consent for the use of all data collected was obtained from participants following the recommendations of the ALSPAC Ethics and Law Committee at the time. Participants can contact the study team at any time to retrospectively withdraw consent for their data to be used. Study participation is voluntary and during all data collection sweeps, information was provided on the intended use of data.

### Measures

#### Built Environment

A latent factor (see **Figure 1**) representing the built environment (explaining 54.6% of variance), previously associated with increased depressive symptoms [3], comprising six observed indicators including: building density, connectivity density, facility richness, walkability, green space and population density. The signs of associations between components of the built environment factor and active travel are likely to be complex. For example, some green spaces may encourage walking [19], yet green space typically decreases as building density increases [20]; at the same time, higher building density often corresponds to greater facility richness [21], which can encourage active travel [22]. Our aim was to represent the built environment more realistically, where facilitators and barriers coexist within the same spatial context. For further information on each of the built environment indicators, see **supplementary section 1.1**. Information on the fit indices of this one factor structure in this ALSPAC sample can be seen in Sanders et al. [3].

**Figure 1.**
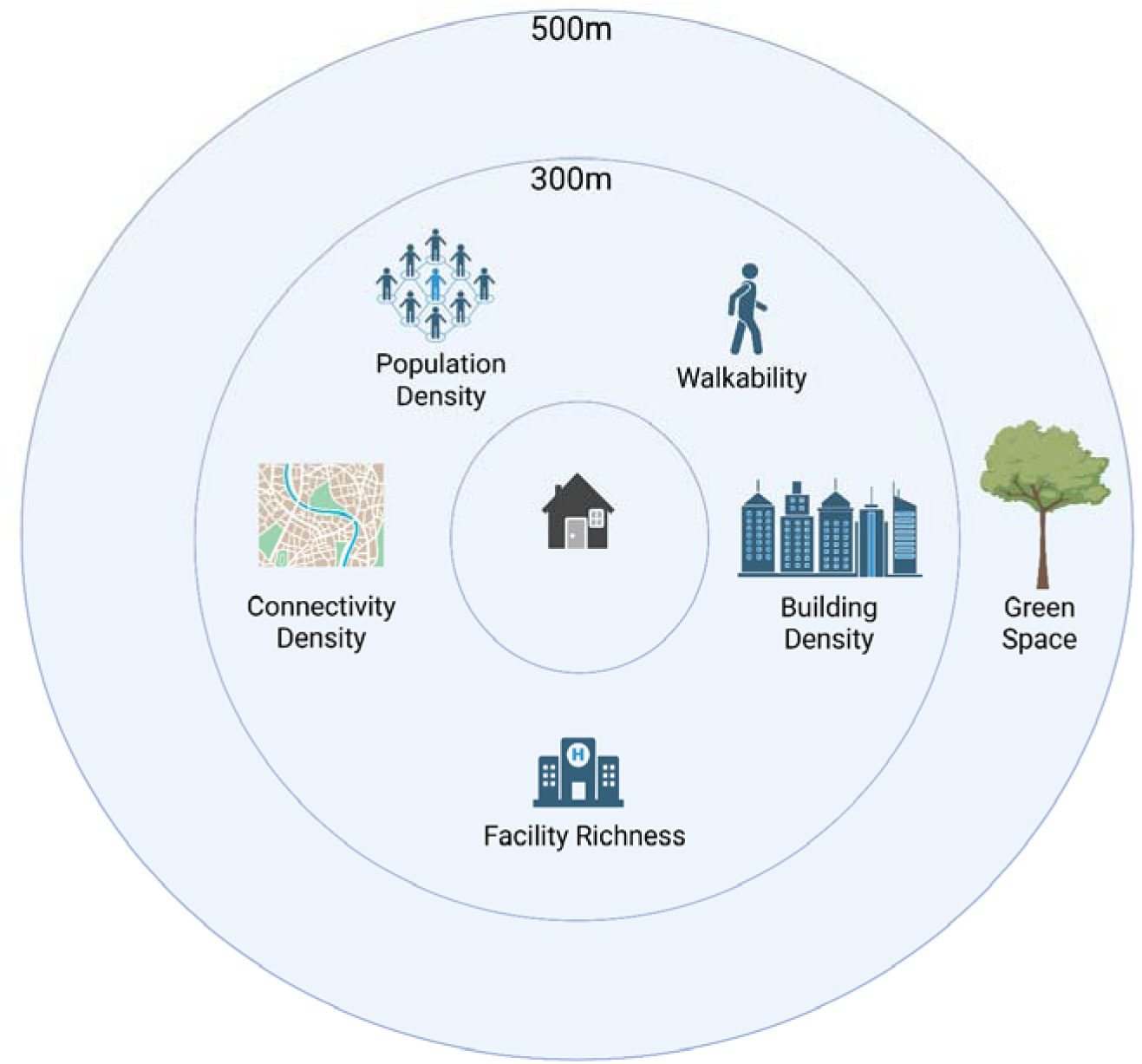
Variables included in the built environment factor used in this study, and the distance buffers at which they were measured.

#### Active Travel Behaviour

At a 2-year follow-up, mothers were asked, ‘How do you usually travel to work?’ with multiple-choice responses including ‘walk’, ‘public transport (bus/train)’, ‘cycle’, ‘car’, ‘work from home’ and ‘other (please describe)’. Active travel behaviours were coded in the following order from lowest to highest active travel scores: (0) Work from home, don’t work, or by car only or car and public transport or other, (1) by public transport or other, (2) walking or cycling and another mode and (3) walking and/or cycling only. At the same time, mothers were also asked, ‘How long does it usually take to travel to work?’ with responses including ‘less than 15 minutes’, ‘15-29 minutes’, ‘30-59 minutes’ or ‘hour or more’. The same question was asked for journey times back from work. In order to make findings more translatable, we took the midpoint of these times and rounded up to the nearest minute. For example, participants who responded with ‘less than 15 minutes’ were given a journey time of 8 minutes, and participants who responded with ‘15-29 minutes’ were given a journey time of 22 minutes. For the ‘hour or more’ category where no maximum is given, 60 minutes was used instead. To better capture the overall dose of active travel exposure, we derived a composite measure by multiplying the ordinal active travel score by total commuting time. This approach integrates both the intensity of travel mode and duration of exposure, consistent with dose-based conceptualisations of physical activity (e.g., MET-minutes [23]), and allows differentiation between individuals with similar travel modes but differing journey lengths.

#### Depressive Symptoms

Depressive symptoms were measured at study enrolment and at a 4-year follow-up using the Crown Crisp Experiential Index (CCEI) [24]. The CCEI depression subscale consisted of 8 items such as, ‘Feels life is too much effort’ and ‘Feels sad for a long time’, with responses including ‘Very often’, ‘Often’, ‘Not very often’ and ‘Never’. Scores can range from 0 to 16. Depressive symptoms were also measured at an 18-year follow-up, using the 36-Item Short Form Survey (SF-36) Mental Health subscale, a valid and reliable measure of mental health [25] and a strong predictor of depression diagnosis [26].

#### Covariates

Covariates included socioeconomic status (SES) risk factors (including education, experience of reduced income, losing a job, becoming homeless, and financial difficulties), baseline depressive symptoms, familial depressive history, physical activity, age at enrolment and depressive symptoms, and smoking (**supplementary section 1.2**).

## Statistical Analysis

This study was preregistered prior to analysis on the Open Science Framework (https://osf.io/exfmd/files/vb7ye) with deviation from the preregistration clearly highlighted. Missing data was imputed. See **supplementary section 1.3** for further information and for a comparison between original and imputed data (**Supplementary Table S1**).

We tested whether the association between the built environment factor (at study enrolment) and depressive symptoms (at 4-year follow-up), identified in our previous work [3], was moderated by active travel behaviour (at 2-year follow-up), when controlling for other known depression predictors.

We performed a pooled moderation analysis on all imputed datasets, testing whether active travel behaviour (at 2-year follow-up) moderates the association between built environment factor (at study enrolment) and depressive symptoms (at 4-year follow-up), whilst adjusting for SES risk factors, baseline depressive symptoms, physical activity, familial depressive history, smoking and age at study enrolment. Moderation was examined using linear regression models including an interaction term between the built environment factor and active travel behaviour. Predictors were mean-centred prior to analysis. We used R Studio Statistical Software (v4.1.0; R Core Team, 2021).

Due to high missingness in active travel behaviour, we also performed a sensitivity analysis restricted to individuals with complete data on active travel behaviour.

## Results

### Descriptive Statistics

Participants were women with a mean age of 28.11 years at study enrolment (**Table 1**). The majority of participants were classified in the car/non-travel group (89.7%), with relatively few engaging in active travel, either exclusively (1.1%) or in combination with other modes (8.5%) for commuting. Shorter journey times were most common, with 19.7% reporting approximately 8 minutes and 12.3% reporting 22 minutes. Please see **Supplementary Table S2** for a full breakdown of active travel categories.

**Table 1.**
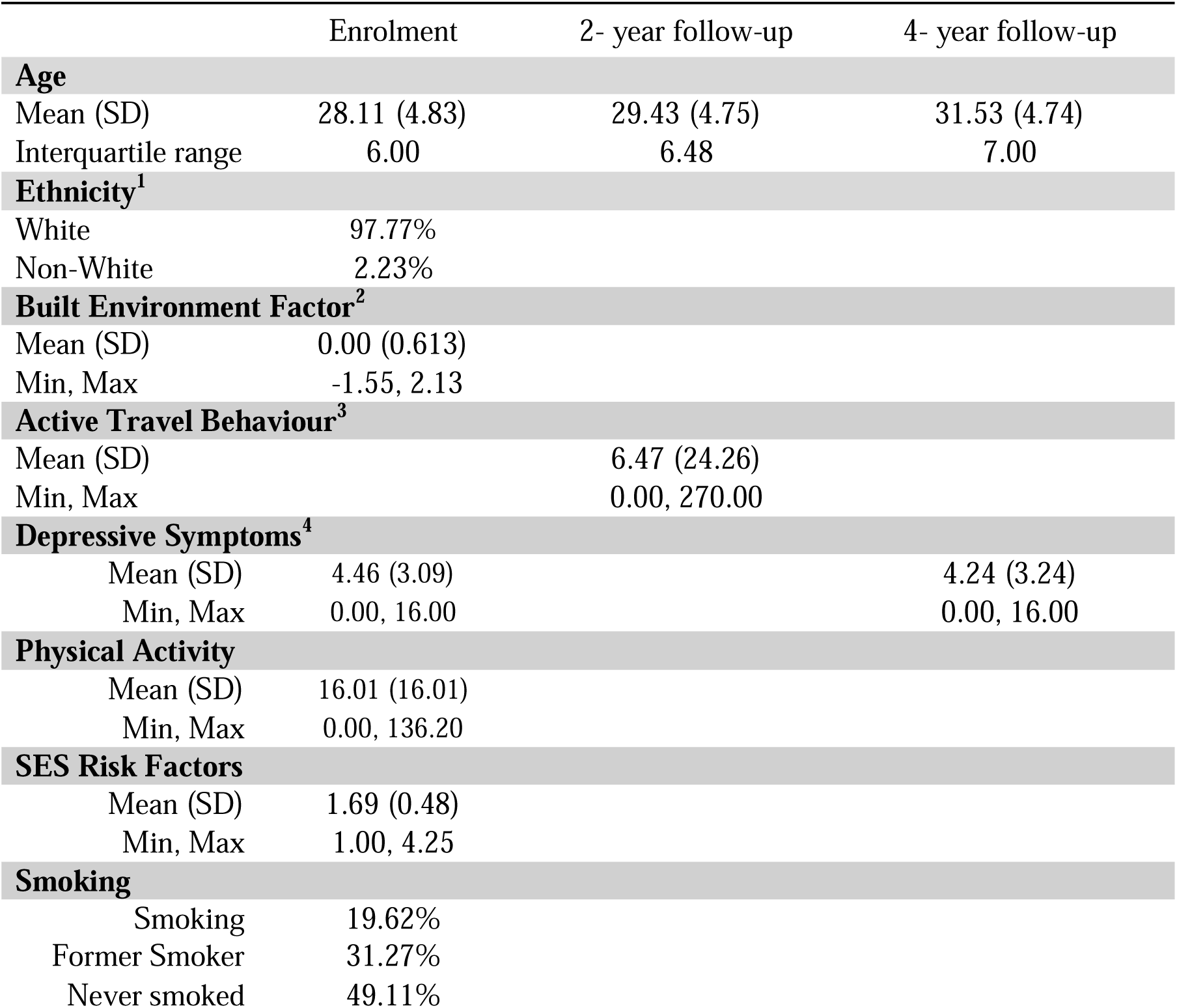
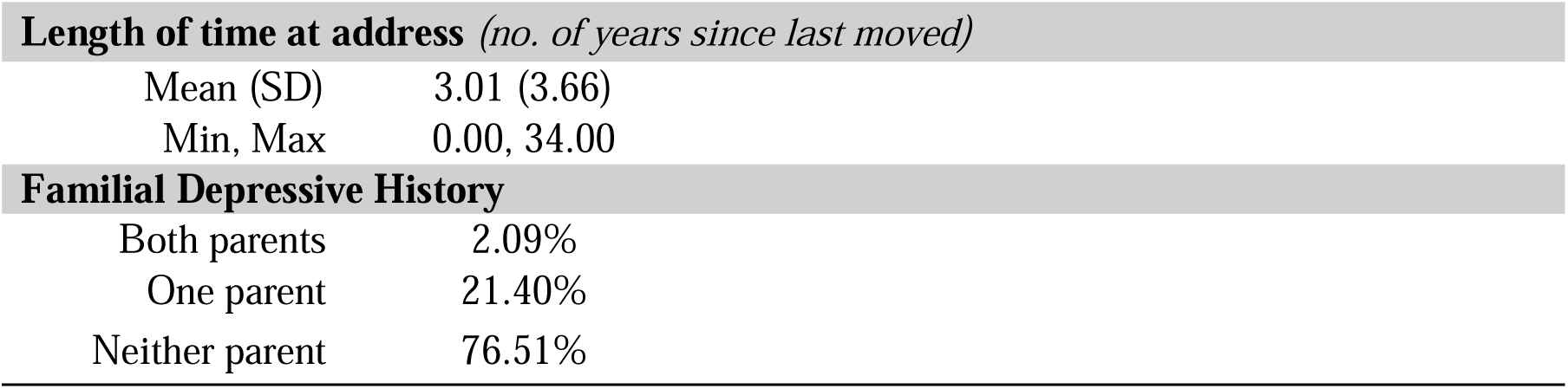
Study descriptives for 10,310 women who had completed questionnaire items for their depressive symptoms at enrolment and/or at the ∼4-year follow-up (imputed data). *^1^ Ethnicity (e.g., white, black/Caribbean, black/African, black/other, Indian, Pakistani, Bangladeshi, Chinese, any other ethnic group) was self-reported at study enrolment*. *^2^ For a breakdown of descriptive statistics of components of the built environment factor, see* ***Supplementary Table S3***. *^3^ Active travel behaviour was calculated by multiplying the ordinal active travel score by total commuting time, derived from the midpoint of categorical journey-time bands (‘less than 15 minutes’, ‘15-29 minutes’, ‘30-59 minutes’ or ‘hour or more’). 60 minutes was used for the hour or more category*. *^4^ Depressive symptoms measured at enrolment and 4-year follow-up used the Crown Crisp Experiential Index*. *See* ***Supplementary Table S4*** *for descriptive statistics of the sensitivity sample (complete active travel data)*. *SD = standard deviation; SES = socioeconomic status*.

The built environment factor was weakly and positively correlated with depressive symptoms at study enrolment (*r* = 0.05, *p* < .001) and the 4-year follow-up (*r* = 0.04, *p* < .001) (**Figure S1**). Active travel behaviour was weakly and positively correlated with the built environment factor (*r* = 0.09, *p* < .001), and weakly and negatively correlated with depressive symptoms at study enrolment (*r* = −0.01, *p* = .129) and the 4-year follow-up (*r* = - 0.01, *p* = .493), yet two of these three correlations were not significant. Depressive symptoms at study enrolment were moderately correlated with symptoms at the 4-year follow-up (*r* = 0.42, *p* < .001).

As shown in **Table 2**, there was no main effect of active travel behaviour (at 2-year follow-up) on depressive symptoms (at 4-year follow-up) (β = −0.002, *p* = .854) in covariate adjusted analyses. In addition, active travel behaviour (at 2-year follow-up) did not moderate the association between the built environment (at study enrolment) and depressive symptoms (at 4-year follow-up) (β = −0.002, *p* = .865). Moreover, the built environment factor did not associate with active travel behaviour in adjusted models (**Figure 2** and **Table 2**). Results were generally consistent in covariate-unadjusted models and in sensitivity analyses, restricted to individuals with complete data on active travel behaviour (**SM Tables S4-S8**). When dichotomising active travel behaviour in further exploratory sensitivity analyses, participants living in areas characterised by higher built environment factor scores had greater odds of engaging in active travel behaviour (**SM Tables S9-S11**). However, as before active travel behaviour did not moderate the association between the built environment factor and depressive symptoms.

**Table 2.**
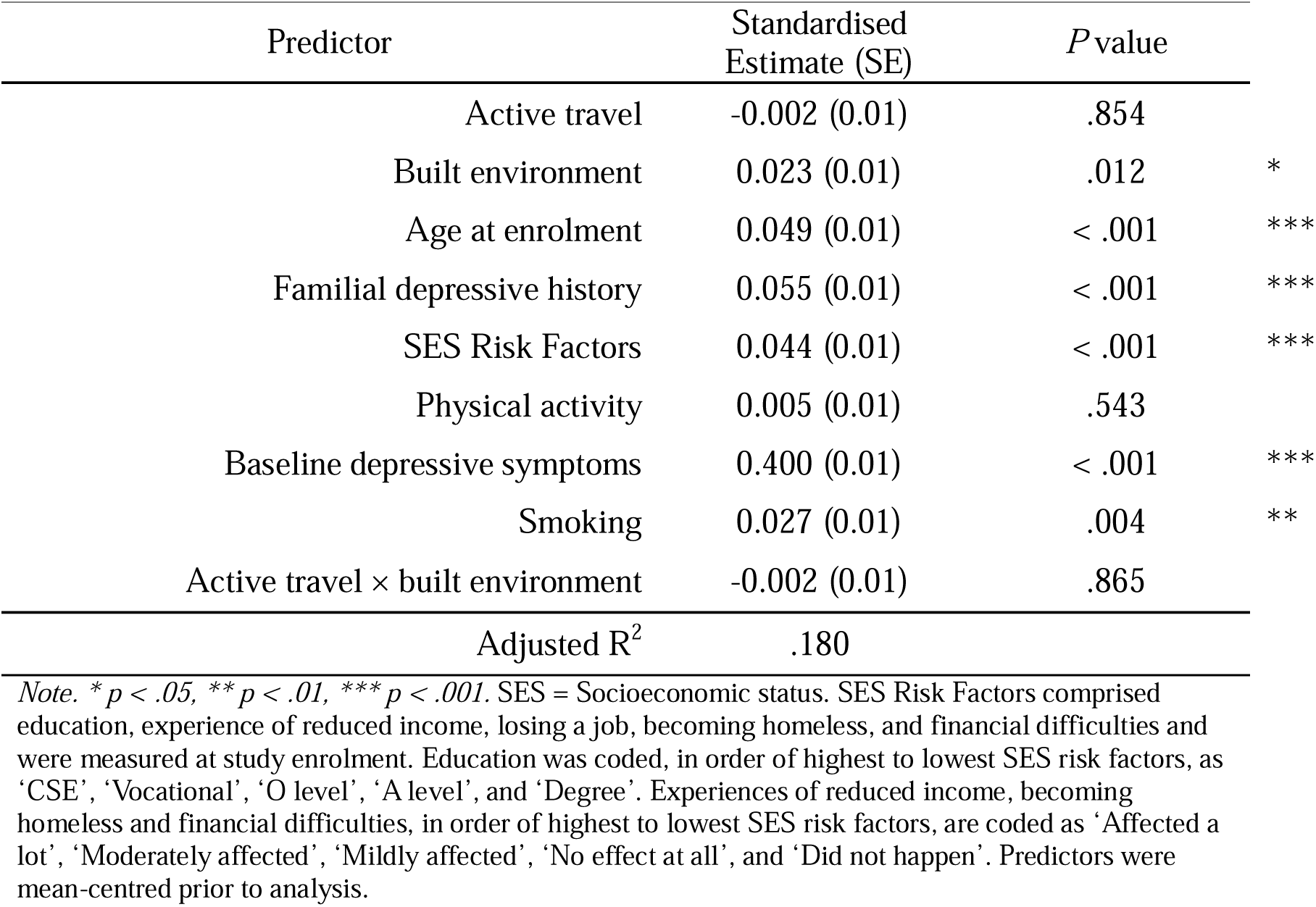
Covariate-adjusted moderation model examining the association between the built environment at enrolment, active travel at 2-year follow-up, and depressive symptoms at 4-year follow-up.

| Predictor | Standardised Estimate (SE) | P value |  |
| --- | --- | --- | --- |
| Active travel | -0.002 (0.01) | .854 |  |
| Built environment | 0.023 (0.01) | .012 | * |
| Age at enrolment | 0.049 (0.01) | < .001 | *** |
| Familial depressive history | 0.055 (0.01) | < .001 | *** |
| SES Risk Factors | 0.044 (0.01) | < .001 | *** |
| Physical activity | 0.005 (0.01) | .543 |  |
| Baseline depressive symptoms | 0.400 (0.01) | < .001 | *** |
| Smoking | 0.027 (0.01) | .004 | ** |
| Active travel × built environment | -0.002 (0.01) | .865 |  |
| Adjusted R<sup>2</sup> | .180 |  |  |

*Note.* \* $p < .05$ , \*\* $p < .01$ , \*\*\* $p < .001$ . SES = Socioeconomic status. SES Risk Factors comprised education, experience of reduced income, losing a job, becoming homeless, and financial difficulties and were measured at study enrolment. Education was coded, in order of highest to lowest SES risk factors, as 'CSE', 'Vocational', 'O level', 'A level', and 'Degree'. Experiences of reduced income, becoming homeless and financial difficulties, in order of highest to lowest SES risk factors, are coded as 'Affected a lot', 'Moderately affected', 'Mildly affected', 'No effect at all', and 'Did not happen'. Predictors were mean-centred prior to analysis.

**Figure 2.**
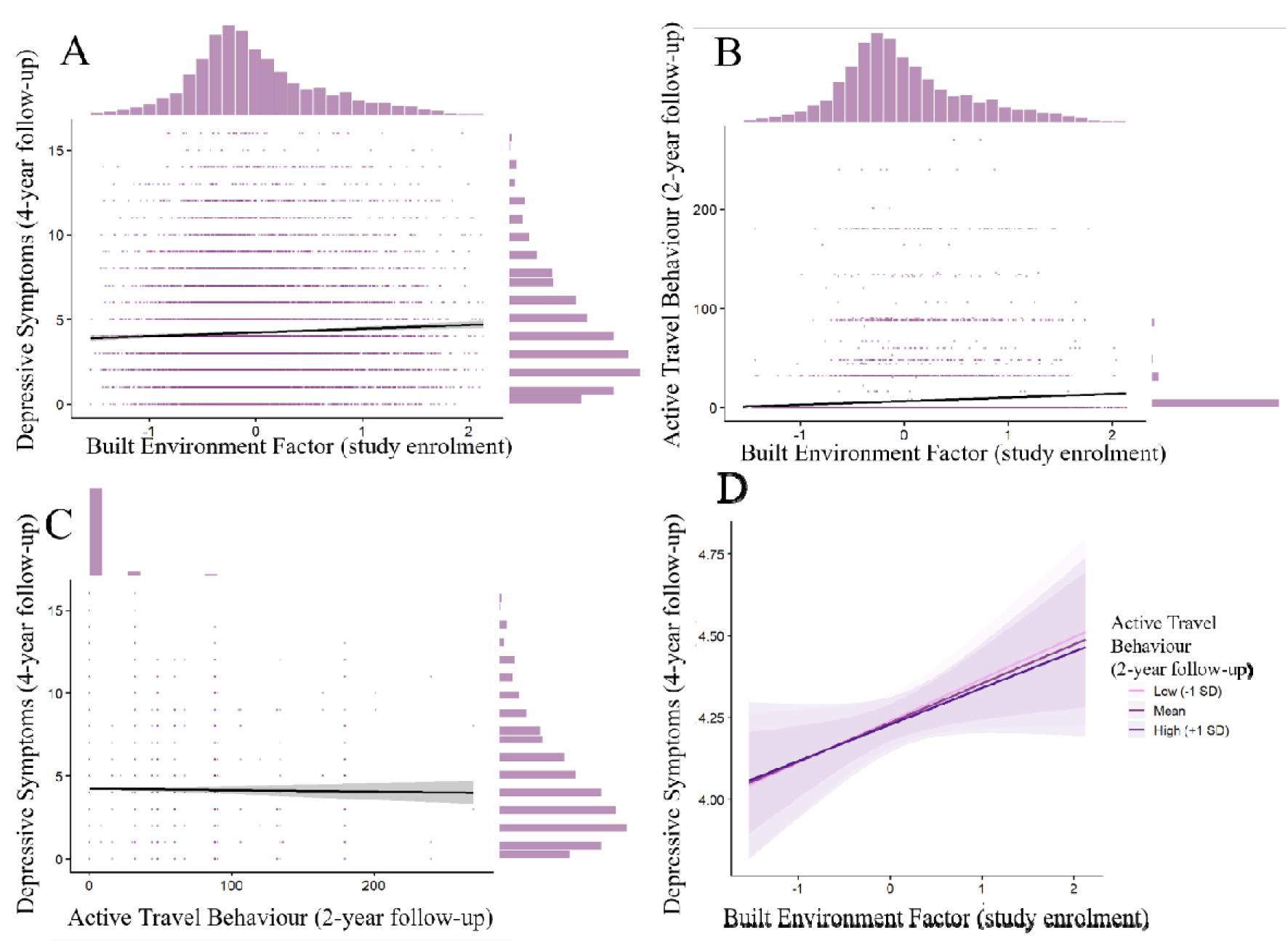
Scatterplots of A) the built environment factor with depressive symptoms; B) the built environment factor with active travel behaviour; and C) active travel behaviour with depressive symptoms. D) Interaction plot showing active travel behaviour and depressive symptoms by built environment level. Please note that the scale for depressive symptoms in panel D is narrower than in the other panels to more clearly display the interaction lines. Active travel behaviour was calculated by multiplying the ordinal active travel score by total commuting time, derived from categorical journey-time bands. Depressive symptoms were measured using the Crown Crisp Experiential Index.

Exploratory analyses examining correlations between individual built environment indicators, active travel behaviour and depressive symptoms showed generally weak associations (**Supplementary Table S12**).

## Discussion

This study aimed to investigate whether the association between a built environment factor and depressive symptoms is moderated by active travel behaviour. Our results confirm the relationship between built environment and depression, but show no evidence to support a moderating role of active travel behaviour in the relationship between a built environment factor and depressive symptoms. We highlight and discuss three key points.

Firstly, our results showed no evidence of an association between active travel behaviour at a 2-year follow-up and depressive symptoms at a 4-year follow-up. This is inconsistent with literature identifying improvements to mental health following active travel workplace interventions [14,27,28]. However, much of this literature has focused on subjective wellbeing rather than symptoms of mental ill-health [28]. However, Mutrie and colleagues [27] did use a well-validated mental health survey. Therefore, this evidence may show that structured and supported active travel interventions confer benefits beyond the travel behaviour itself. For example, intervention components such as activity monitoring, goal setting, or the provision of safety information may contribute to improved psychological wellbeing by enhancing motivation, self-efficacy, or perceived safety during commuting. Furthermore, participants in some interventions were recruited based on an existing interest in active travel [27]. Consequently, observed mental health improvements may have reflected the achievement of personally meaningful behavioural goals, rather than the effects of active travel alone.

A lack of association between active travel behaviour at a 2-year follow-up and depressive symptoms at a 4-year follow-up is also inconsistent with a large body of literature identifying protective effects of general physical activity on mental health [29]. Therefore, it may also be that the active travel behaviour measured in this study did not reach a threshold of intensity or duration to see protective effects of physical activity on depressive symptoms, which is generally consistent with research identifying active travel commuting methods representing the lower range of intensity of exercise [30].

Secondly, despite observing a weakly positive correlation between the built environment factor and active travel behaviour, the association weakened after covariate adjustment. Previous research has found features within the built environment, including walkability and supportive transport infrastructure, to be strong predictors of active travel behaviour [10,11]. However, the built environment factor used in this study encompassed a broader range of environmental features, including green space and building density. As such, individual variability in their associations with active travel behaviour could have weakened the overall association with the built environment factor. However, exploratory analyses evidenced consistent results, whereby weak positive correlations were generally observed between the individual built environment features with active travel behaviour, providing evidence to the contrary. Nevertheless, our sensitivity analyses where active travel behaviour was dichotomised, did show evidence of an association between the built environment factor and whether people engage in active travel behaviour or not. Therefore, our findings may suggest the built environment is more strongly associated with whether individuals engage in active travel at all, rather than the amount undertaken.

Thirdly, contrary to our hypothesis, we did not see any evidence to support a moderating effect of active travel behaviour on the association between the built environment factor and depressive symptoms. This means the association between the built environment factor and depressive symptoms did not differ depending on active travel behaviour. We hypothesised that associations between the built environment factor and depressive symptoms would be weaker in individuals who engaged more with active travel. In other words, active travel behaviour would act as a protective buffer over this relationship. Overall, active travel behaviour is thought to have a net positive impact on health [31]. However, active travel behaviour can also expose individuals to pollutants and injury risk, which are more probable in urban environments [31,32]. Consequently, whilst these competing influences may have attenuated any weak beneficial effects of active travel, they are unlikely to fully explain the absence of a moderating effect observed in the present study. Instead, this finding may reflect either a genuinely limited moderating role of active travel behaviour or methodological limitations of the study.

Our findings must be considered alongside limitations. Firstly, there is likely to be considerable heterogeneity within intensity of activity and actual distance travelled within the same categories of both travel mode and duration. Similarly, journey durations may have varied substantially within the predefined duration categories. As a result, these measurement errors may have attenuated any associations between active travel behaviour and either the built environment factor or depressive symptoms. Moreover, whilst the built environment was measured at study enrolment, active travel behaviour was measured at a 2-year follow-up. Participants may have changed residence during this period, resulting in active travel behaviours being shaped by different environmental contexts than those captured by the built environment measure. Perhaps most notably, active travel behaviour was assessed during a period when the participants were caring for young infants, a period in which physical activity and travel patterns often differ from those with older children and no children [33,34]. As a result, levels of active travel reported may not accurately reflect participants’ typical travel behaviours over the longer term. Relatedly, active travel behaviour was measured in this study only in the context of commuting behaviour, and not all trip purposes. Whilst evidence from the 1990s suggests up to 73% of employed mothers in the UK returned to work within 12 months [35], this percentage is likely slightly lower in the ALSPAC sample, where 67.2% of mothers had returned to work by 34 months postpartum [36]. As a result, the measurement of active travel in this study likely underestimated the actual level of active travel engagement in the sample. Given that depressive symptoms were measured years later, active travel behaviour may have had limited relevance to subsequent mental health outcomes, potentially weakening observed associations. Finally, active travel behaviour was relatively uncommon within the cohort, potentially limiting variability in the moderator and reducing statistical power to detect interaction effects. However, it is notable that no evidence of moderation was observed even when active travel was operationalised as a binary exposure in sensitivity analyses, suggesting that limited power alone is unlikely to explain the null interaction findings.

To conclude, this study set out to investigate whether active travel behaviour at a 2-year follow-up moderated the association between the built environment factor at study enrolment and depressive symptoms at a 4-year follow-up. Our evidence suggests that whilst there is an association between a built environment factor and depressive symptoms, there was no evidence to support this was moderated by active travel behaviour. Hence, this study provides no support that engagement in active travel behaviour would be an effective intervention to mitigate mental-health related risks of living in urban environments. Future research may wish to explore these relationships in different segments of the population (including men and older adults) and across different active travel profiles to better understand whether these interventions may be effective outside of this study population.

## Availability of data and materials

The code used to perform the analyses in this study is publicly available on GitHub at: https://github.com/fsanders1/active-travel-moderation. The informed consent obtained from ALSPAC (Avon Longitudinal Study of Parents and Children) participants does not allow the data to be made available through any third party maintained public repository. Supporting data are available from ALSPAC on request under the approved proposal number, B2957. Full instructions for applying for data access can be found here: http://www.bristol.ac.uk/alspac/researchers/access/.

## Authors’ contributions

F.S: Conceptualisation, analysis, writing original draft, reviewing and editing. V.B.: conceptualisation, reviewing and editing. L.W.: conceptualisation, reviewing and editing. E.W: conceptualisation, supervision, project administration, acquisition of funding, reviewing and editing.

## Acknowledgements

We are extremely grateful to all the families who took part in this study, the midwives for their help in recruiting them, and the whole ALSPAC team, which includes data collection staff, data and administrations staff, technical managers and the technical staff with the Bristol Bioresource Laboratory, based within the University of Bristol.

