## Supplementary Materials for "Does active travel behaviour moderate the association between the built environment and depression?"

**Supplementary Section 1.1**

*Built Environment Factor Indicators*

- Building Density: Building density is measured at study enrolment and is measured by the number of m^2^ of buildings per km^2^. A 300m buffer score is used which represents the average building density within a 300m radius of the participants’ home address. A 300m buffer is used for most built environment variables as it is the more consistently used measure in literature, and there are high correlations amongst different distance buffers and health behaviours [1–4]. A 300m buffer hence strikes a balance between capturing a sufficient portion of the built environment with a strong likelihood of the participant interacting with that portion of their environment. Building density showed a moderate loading on the built environment factor (λ = 0.65), indicating a substantial contribution to the association with increased depressive symptoms.
- Connectivity Density: Connectivity density at a 300m buffer is measured at study enrolment. This is characterised as the number of road intersections per km^2^. Road intersections are places where two or more roads connect in some way, either through meeting or diverging [5]. The number of intersections has been linked to increases in walking behaviour [6], and loaded strongly onto the built environment factor (λ = 0.80), suggesting it is a key contributor to the association with increased depressive symptoms.
- Facility Richness: Facility richness at a 300m buffer is measured at study enrolment. Facility richness is measured as the number of different facility types in the 300m buffer (e.g., educational, health) available divided by the maximum potential number of facility types. It is defined through a score of 0 to 1, where greater scores indicate greater facility richness. Greater facility richness at residential addresses has been associated with increased walking behaviour [6], and also loaded strongly on the built environment factor (λ = 0.81) associated with increased depressive symptoms.
- Walkability: Walkability is measured at study enrolment and is measured as an average walkability index score (0-1) within a 300m radius of participants’ home addresses. The walkability score is defined as the mean of deciles for facility richness index, connectivity density, land use diversity and population density [7,8]. The walkability index contained some components of built environment features included in the analysis, such as facility richness, population density and connectivity density. However, we used the walkability score in addition to these components for two reasons. Firstly, it is recommended by Descarpentrie and colleagues [3] to use both the walkability scores and the component variables (facility richness index etc.) in order to capture any potential information losses that can occur from transforming data into a single composite score. Secondly, including walkability scores are also important because not all components of walkability are included individually in this analysis, including pedestrian infrastructure and traffic safety measures. Greater scores of walkability are strongly associated with increased walking behaviour [9]. Walkability also showed the strongest loading on the built environment factor (λ = 0.87) associated with increased depressive symptoms.
- Green Space: Green space is measured at study enrolment and was quantified using the Normalized Difference Vegetation Index at a 500m buffer. The 500m buffer is selected as this is the maximum buffer radius available, and greater NDVI buffer sizes have been found to have stronger associations with health than smaller sizes [10]. Greater buffer sizes are also important as individuals often report travelling substantially further from their home addresses specifically to reach higher quality green spaces, particularly for recreational trips [11]. Scores range from -1 to +1, with greater scores indicating more green space. Greater green space has been previously associated with increased walking [12] and loaded strongly and negatively onto the built environment factor (λ = −0.74) associated with increased depressive symptoms.
- Population Density: Population density is measured at study enrolment and is defined through the number of inhabitants per km^2^. High density residential areas have been associated with increased walking behaviour [6] and population density showed a moderate loading on the built environment factor (λ = 0.51) associated with increased depressive symptoms.

**Supplementary Section 1.2**

*Covariates*

Socioeconomic Status Risk Factors: SES risk factors scores – comprise mother’s education, experience of reduced income, losing a job, becoming homeless, and financial difficulties – are measured at study enrolment. Education is coded, in order of highest to lowest SES risk factors, as ‘CSE’, ‘Vocational’, ‘O level’, ‘A level’, and ‘Degree’. Experiences of reduced income, becoming homeless and financial difficulties, in order of highest to lowest SES risk factors, are coded as ‘Affected a lot’, ‘Moderately affected’, ‘Mildly affected’, ‘No effect at all’, and ‘Did not happen’. These are measured at study enrolment. SES is important to control for as it is associated with depressive symptoms, urban living and active travel behaviour [13–15].

Familial Depressive History: Familial depressive history (e.g., known lifetime experience of depressive symptoms or consistent low mood in the participants’ parents) is reported by participants at study enrolment (2=both parents, 1=one parent, 0=none). Controlling for family history of depression is important because it is a key predictor of depressive symptoms [16].

Smoking: Smoking is self-reported at study enrolment, and responses measured whether participants had never smoked, were a former smoker, or were still currently smoking. Smoking is important to control for as it associates with both depressive symptoms and urban living [17,18].

Physical activity: At study enrolment, mothers were asked ‘How much do you do the following at present?’ followed by ‘jogging’, ‘aerobics’, ‘ante natal exercise’, ‘keep fit’, ‘yoga’, ‘squash’, ‘tennis/badminton’, ‘swimming’, ‘brisk walking’, ‘weight training’, ‘cycling’ ‘other exercise’. Mothers could then respond with, ‘Never’, ‘Less than one hour per week’, ‘2-6 hours per week’, and ‘more than 7 hours per week’. As described by Liu and colleagues (2011), scores were then weighted according to average metabolic equivalent scores meaning they represented the weekly total energy expenditure from activities reported. Higher scores represent greater total energy expenditure from reported physical activities. Controlling for general levels of physical activity is important because it associates with depressive symptoms and active travel behaviour [19,20].

Age at Study Enrolment: ALSPAC built environment data was collated from multiple external sources and then retrospectively added to their database using participant addresses. Hence, similarly to other researchers [21], we are using mothers’ age at study enrolment to capture the age at built environment as they were measured within the same 12-month window. Controlling for age is important as it associates with both depressive symptoms and urban living [22,23].

**Supplementary Section 1.3**

*Missing Data Imputation*

As part of a previous study [24], imputation of missing data had already been performed. We had previously selected mothers who had at least 50% data available for built environment features (including building density (300m buffer), connectivity density (300m buffer), facility density (300m buffer), facility richness (300m buffer), bus lines length (300m buffer), bus stops (500m buffer), total traffic load (100m buffer), distance to nearest road, walkability (300m buffer), green space (500m buffer) and population density) and depressive symptoms at study enrolment, leaving up to 10,310 mothers available for analysis. We originally intended to also include bus stops and traffic load as built environment features, but this was not possible due to too much missing data (99.5% and 91.5% respectively). The MICE R package [25] was used to impute missing values, through Random Forest, in built environment (at study enrolment) and depressive symptoms (only at follow-ups) variables, active travel behaviour, as well as covariates (including maternal age at enrolment, physical activity, familial depressive history, length of time at address, smoking, SES risk factors). Participants were removed if they had more than 50% missingness in built environment variables and no depressive symptom data at study enrolment. Built environment features, active travel behaviour, depressive symptoms at all time points and all covariates were used as predictors in a predictor matrix to impute missing data. Auxiliary variables (i.e., variables selected to inform imputation but not used in the analysis) included depressive symptoms at an 18-year follow-up, child ethnicity (at age 17), child sex, child travel behaviour at age 13 (and age at measurement), child physical health (at age 13), child pro-environmentalism (at age 13), and child cognition at age 24 (and age at measurement). Auxiliary variables are variables which are going to be used in later analyses, have been identified as possible moderators or are closely associated with variables in this study in order to reduce bias [26].

Imputation was performed using 60 iterations and 30 imputed datasets. The predictor matrix was specified using quickpred(), with a minimum correlation set to 0.05, striking a balance between having sufficient variable breadth yet avoiding weak predictors.

Imputation was performed at the item level, and variables including active travel behaviour were constructed after imputation.

**Table S1.**

*Original data compared to pooled (average across MICE datasets) imputed data (30 imputations).*

**For age, the 25th and 75th percentiles are reported instead of minimum and maximum values.*

|  | **N** | | **Mean** | | **SD** | | **Min** | | **Max** | | **Difference Statistic** | ***P* Value** |
| --- | --- | --- | --- | --- | --- | --- | --- | --- | --- | --- | --- | --- |
|  | Original | Imputed | Original | Imputed | Original | Imputed | Original | Imputed | Original | Imputed |  |  |
| Age at enrolment* | 10310 | 10310 | 28.11 | 28.11 | 4.83 | 4.83 | 25.00 | 25.00 | 31.00 | 31.00 | t = 0.00 | .99 |
| Building Density | 10310 | 10310 | 400733.27 | 400733.27 | 135306.01 | 135306.01 | 20000.00 | 20000.00 | 670000.00 | 670000.00 | t = 0.00 | .99 |
| Connectivity Density | 10310 | 10310 | 129.73 | 129.73 | 60.19 | 60.19 | 10.00 | 10.00 | 340.00 | 340.00 | t = 0.00 | .99 |
| Facility Richness | 10310 | 10310 | 0.07 | 0.07 | 0.07 | 0.07 | 0.00 | 0.00 | 0.30 | 0.30 | t = 0.00 | .99 |
| Green Space | 10310 | 10310 | 0.42 | 0.42 | 0.09 | 0.09 | 0.20 | 0.20 | 0.70 | 0.70 | t = 0.00 | .99 |
| Population density | 10169 | 10310 | 4915.82 | 4870.93 | 2740.44 | 2752.60 | 1000.00 | 1000.00 | 10000.00 | 10000.00 | t = 1.13 | .26 |
| Walkability | 10072 | 10310 | 0.30 | 0.30 | 0.06 | 0.06 | 0.15 | 0.15 | 0.50 | 0.50 | t = 1.12 | .27 |
| Travel modes | | | | | | | | | | | | |
| Car | | | | | | | | | | | | |
| N | 9768 | 10310 | NA | | | | | | | | χ² = 123.31 | < .001 |
| Yes | 38.16% | 31.36% |  |  |  |  |  |  |  |  |  |  |
| No | 61.84% | 68.64% |  |  |  |  |  |  |  |  |  |  |
| Public transport | | | | | | | | | | | | |
| N | 7848 | 10310 | NA | | | | | | | | χ² = 13.04 | .0003 |
| Yes | 6.98% | 5.76% |  |  |  |  |  |  |  |  |  |  |
| No | 93.02% | 94.24% |  |  |  |  |  |  |  |  |  |  |
| Cycling | | | | | | | | | | | | |
| N | 7573 | 10310 | NA | | | | | | | | χ² = 2.36 | .125 |
| Yes | 1.44% | 1.19% |  |  |  |  |  |  |  |  |  |  |
| No | 98.56% | 98.81% |  |  |  |  |  |  |  |  |  |  |
| Walking | | | | | | | | | | | | |
| N | 7862 | 10310 | NA | | | | | | | | χ² = 4.97 | .026 |
| Yes | 8.18% | 7.35% |  |  |  |  |  |  |  |  |  |  |
| No | 91.82% | 92.65% |  |  |  |  |  |  |  |  |  |  |
| Other | | | | | | | | | | | | |
| N | 7176 | 10310 | NA | | | | | | | | χ² = 0.47 | .468 |
| Yes | 1.03% | 0.92% |  |  |  |  |  |  |  |  |  |  |
| No | 98.97% | 99.08% |  |  |  |  |  |  |  |  |  |  |
| Works from home | | | | | | | | | | | | |
| N | 10246 | 10310 | NA | | | | | | | | χ² = 20.92 | < .001 |
| Yes | 3.76% | 2.74% |  |  |  |  |  |  |  |  |  |  |
| No | 96.24% | 97.26% |  |  |  |  |  |  |  |  |  |  |
| Does not work | | | | | | | | | | | | |
| N | 10246 | 10310 | NA | | | | | | | | χ² = 325.47 | < .001 |
| Yes | 52.71% | 63.98% |  |  |  |  |  |  |  |  |  |  |
| No | 47.29% | 36.02% |  |  |  |  |  |  |  |  |  |  |
| Outward Journey Duration | | | | | | | | | | | | |
| N | 10165 | 10310 | NA | | | | | | | | χ² = 271.31 | < .001 |
| 0 minutes | 56.92% | 62.90% |  |  |  |  |  |  |  |  |  |  |
| 8 minutes | 21.82% | 19.70% |  |  |  |  |  |  |  |  |  |  |
| 22 minutes | 14.85% | 12.30% |  |  |  |  |  |  |  |  |  |  |
| 45 minutes | 5.61% | 4.60% |  |  |  |  |  |  |  |  |  |  |
| 60 minutes | 0.80% | 0.60% |  |  |  |  |  |  |  |  |  |  |
| Return Journey Duration | | | | | | | | | | | | |
| N | 9786 | 10310 | NA | | | | | | | | χ² = 167.59 | < .001 |
| 0 minutes | 59.13% | 62.90% |  |  |  |  |  |  |  |  |  |  |
| 8 minutes | 20.74% | 19.90% |  |  |  |  |  |  |  |  |  |  |
| 22 minutes | 13.71% | 11.70% |  |  |  |  |  |  |  |  |  |  |
| 45 minutes | 5.63% | 4.80% |  |  |  |  |  |  |  |  |  |  |
| 60 minutes | 0.79% | 0.60% |  |  |  |  |  |  |  |  |  |  |
| Familial Depressive History | | | | | | | | | | | | |
| N | 10310 | 10310 | NA | | | | | | | | χ² = 0.00 | .99 |
| Both parents | 2.09% | 2.09% |  |  |  |  |  |  |  |  |  |  |
| One parent | 21.41% | 21.41% |  |  |  |  |  |  |  |  |  |  |
| Neither parents | 76.51% | 76.51% |  |  |  |  |  |  |  |  |  |  |
| Ethnicity | | | | | | | | | | | | |
| N | 9500 | 10310 | NA | | | | | | | | χ² = 0.36 | .55 |
| White | 97.62% | 97.77% |  |  |  |  |  |  |  |  |  |  |
| Non-White | 2.38% | 2.23% |  |  |  |  |  |  |  |  |  |  |
| Smoking (study enrolment) | | | | | | | | | | | | |
| N | 9063 | 10310 | NA | | | | | | | | χ² = 4.71 | .09 |
| Smoking | 19.17% | 19.62% |  |  |  |  |  |  |  |  |  |  |
| Former smoker | 30.21% | 31.27% |  |  |  |  |  |  |  |  |  |  |
| Never smoked | 50.62% | 49.11% |  |  |  |  |  |  |  |  |  |  |
| Length of time at address | 9446 | 10310 | 3.03 | 3.01 | 3.64 | 3.66 | 0.00 | 0.00 | 34.00 | 34.23 | t = 0.43 | .67 |
| Physical Activity | 10254 | 10310 | 16.00 | 16.00 | 16.01 | 16.01 | 0.00 | 0.00 | 136.20 | 136.20 | t = -0.03 | .97 |
| Age at Active Travel* | 7613 | 10310 | 29.94 | 29.43 | 4.64 | 4.75 | 26.35 | 26.00 | 32.19 | 32.00 | t = 7.16 | < .001 |
| SES Risk Factors | 8390 | 10310 | 1.66 | 1.69 | 0.47 | 0.48 | 1.00 | 1.00 | 4.25 | 4.25 | t = -4.60 | < .001 |
| Depressive Symptoms (study enrolment) | 10310 | 10310 | 4.46 | 4.46 | 3.09 | 3.09 | 0.00 | 0.00 | 16.00 | 16.00 | t = 0.00 | .99 |
| Depressive symptoms (4-year follow-up) | 7489 | 10310 | 4.17 | 4.24 | 3.21 | 3.24 | 0.00 | 0.00 | 16.00 | 16.00 | t = -1.39 | .17 |

**Table S2**

*Breakdown of active travel score component variables at 2-year follow-up.*

| Category | N (Percent) |
| --- | --- |
| **Travel Behaviour Category** | |
| Active travel only | 117 (1.1%) |
| Active travel and other modes | 875 (8.5%) |
| Public transport or other only | 72 (0.7%) |
| Car or not working or none | 9246 (89.7%) |
| **Outward journey** | |
| 0 minutes | 6485 (62.9%) |
| 8 minutes | 2029 (19.7%) |
| 22 minutes | 1263 (12.3%) |
| 45 minutes | 473 (4.6%) |
| 60 minutes | 60 (0.6%) |
| **Return journey** | |
| 0 minutes | 6489 (62.9%) |
| 8 minutes | 2056 (19.9%) |
| 22 minutes | 1206 (11.7%) |
| 45 minutes | 499 (4.8%) |
| 60 minutes | 60 (0.6%) |
| *Note. The midpoint of categorical journey-time bands are presented (‘less than 15 minutes’ = 8 minutes, ‘15-29 minutes’ = 22 minutes, ‘30-59 minutes’ = 45 minutes or ‘hour or more’). 60 minutes was used for the hour or more category. Journey time was recorded independently of mode of travel. A journey time of 0 minutes represents participants who were not working or worked from home.* | |

**Table S3**

*Study descriptives for 10,310 women* *who had completed questionnaire items for their depressive symptoms at enrolment and/or at the ~4-year follow-up (imputed data).*
*^1^ Ethnicity (e.g., white, black/Caribbean, black/African, black/other, Indian, Pakistani, Bangladeshi, Chinese, any other ethnic group) was self-reported at study enrolment. ^2^ Active travel behaviour was calculated by multiplying the ordinal active travel score by total commuting time, derived from the midpoint of categorical journey-time bands (‘less than 15 minutes’, ‘15-29 minutes’, ‘30-59 minutes’ or ‘hour or more’). 60 minutes was used for the hour or more category. ^3^ Depressive symptoms measured at enrolment and a 4-year follow-up used the Crown Crisp Experiential Index.*
*SD = standard deviation; SES = socioeconomic status.*

|  | Enrolment | 2-year follow-up | 4- year follow-up |
| --- | --- | --- | --- |
| **Age** | | | |
| Mean (SD) | 28.12 (4.83) | 29.43 (4.75) | 31.53 (4.74) |
| Interquartile range | 6.00 | 6.48 | 7.00 |
| **Ethnicity^1^** | | | |
| White | 97.77% |  |  |
| Non-White | 2.23% |  |  |
| **Built environment** | | | |
| *Facility Richness* *(no. of different facility types in the 300m buffer available, divided by the maximum potential number of facility types)* | | | |
| Mean (SD) | 0.07 (0.07) |  |  |
| Min, Max | 0.00, 0.30 |  |  |
| *Population Density (number of inhabitants per km^2^)* | | | |
| Mean (SD) | 4870.93 (2752.60) |  |  |
| Min, Max | 1,000.00, 10,000.00 |  |  |
| *Green Space (Normalized Difference Vegetation Index, ranging from -1 to +1)* | | | |
| Mean (SD) | 0.42 (0.09) |  |  |
| Min, Max | 0.20, 0.70 |  |  |
| *Walkability* (*average walkability index score (0-1)*) | | | |
| Mean (SD) | 0.30 (0.06) |  |  |
| Min, Max | 0.15, 0.50 |  |  |
| *Building Density* *(no. of m^2^ of buildings per km^2^)* | | | |
| Mean (SD) | 400733.27 (135306.01) |  |  |
| Min, Max | 20,000.00, 670,000.00 |  |  |
| *Connectivity Density* (*no. of road intersections per km^2^*) | | | |
| Mean (SD) | 129.73 (60.19) |  |  |
| Min, Max | 10.00, 340.00 |  |  |
| **Active Travel Behaviour^2^** | | | |
| Mean (SD) |  | 6.47 (24.26) |  |
| Min, Max |  | 0.00, 270.00 |  |
| **Depressive Symptoms^3^** | | | |
| Mean (SD) | 4.46 (3.09) |  | 4.24 (3.24) |
| Min, Max | 0.00, 16.00 |  | 0.00, 16.00 |
| **Physical Activity** | | | |
| Mean (SD) | 16.01 (16.01) |  |  |
| Min, Max | 0.00, 136.20 |  |  |
| **SES Risk Factors** | | | |
| Mean (SD) | 1.69 (0.48) |  |  |
| Min, Max | 1.00, 4.25 |  |  |
| **Smoking** |  |  |  |
| Smoking | 19.62% |  |  |
| Former Smoker | 31.27% |  |  |
| Never smoked | 49.11% |  |  |
| **Length of time at address** *(no. of years since last moved)* | | | |
| Mean (SD) | 3.01 (3.66) |  |  |
| Min, Max | 0.00, 34.00 |  |  |
| **Familial Depressive History** | | | |
| Both parents | 2.09% |  |  |
| One parent | 21.40% |  |  |
| Neither parents | 76.51% |  |  |

**Figure S1**

*Correlation matrix between variables.*

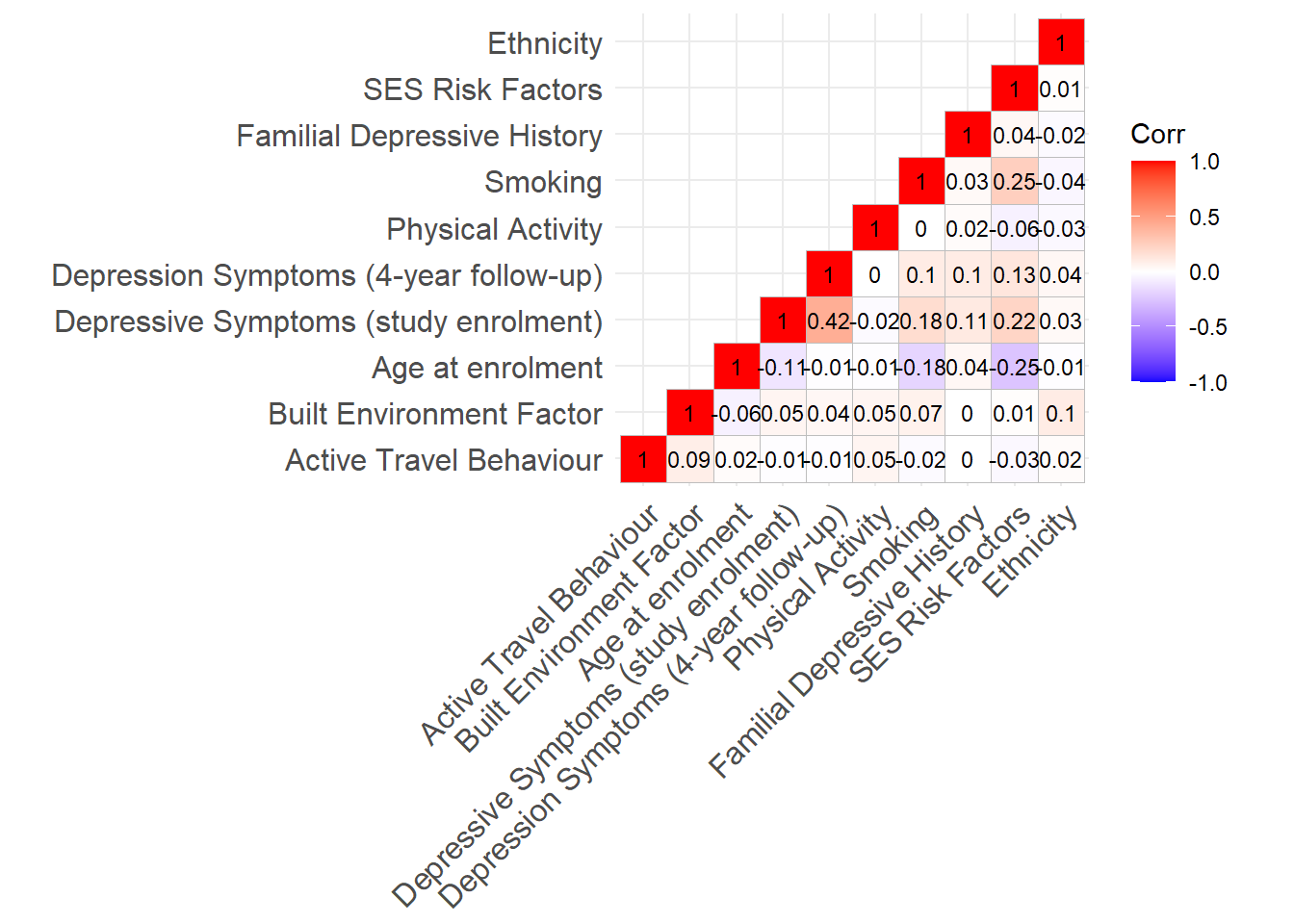

**Table S4**

*Study descriptives for 5,498 women* *in the sensitivity analysis sample* *who had completed questionnaire items for their depressive symptoms at enrolment and/or at the ~4-year follow-up (imputed data) and had complete data for active travel behaviour.*
*^1^ Ethnicity (e.g., white, black/Caribbean, black/African, black/other, Indian, Pakistani, Bangladeshi, Chinese, any other ethnic group) was self-reported at study enrolment. ^2^ Active travel behaviour was calculated by multiplying the ordinal active travel score by total commuting time, derived from the midpoint of categorical journey-time bands (‘less than 15 minutes’, ‘15-29 minutes’,‘30-59 minutes’ or ‘hour or more’). 60 minutes was used for the hour or more category. ^3^ Depressive symptoms measured at enrolment and a 4-year follow-up used the Crown Crisp Experiential Index.*
*SD = standard deviation; SES = socioeconomic status.*

|  | Enrolment | 2-year follow-up | 4-year follow-up |
| --- | --- | --- | --- |
| **Age** | | |  |
| Mean (SD) | 28.41 (4.73) | 29.66 (4.74) | 31.76 (4.71) |
| Interquartile range | 6.00 | 6.00 | 6.00 |
| **Ethnicity^1^** | | |  |
| White | 98.50% |  |  |
| Non-white | 1.50% |  |  |
| **Built Environment Factor** | | |  |
| Mean (SD) | 0.00 (0.62) |  |  |
| Min, Max | -1.55, 2.19 |  |  |
| **Active Travel Behaviour^2^** | | |  |
| Mean (SD) |  | 3.72 (18.91) |  |
| Min, Max |  | 0.00, 270.00 |  |
| **Depressive Symptoms^3^** | | |  |
| Mean (SD) | 4.36 (3.03) |  | 4.25 (3.21) |
| Min, Max | 0.00, 16.00 |  | 0.00, 16.00 |
| **Physical Activity** | | |  |
| Mean (SD) | 16.02 (15.55) |  |  |
| Min, Max | 0.00, 128.60 |  |  |
| **SES Risk Factors** | | |  |
| Mean (SD) | 1.67 (0.48) |  |  |
| Min, Max | 1.00, 4.25 |  |  |
| **Familial Depressive History** | | |  |
| Neither parent | 76.30% |  |  |
| One parent | 21.50% |  |  |
| Both parents | 2.20% |  |  |
| **Smoking** | | |  |
| Never | 50.60% |  |  |
| Until pregnancy | 30.50% |  |  |
| During pregnancy | 18.90% |  |  |

In **Table S5**, we can see the differences between the analytic sample and the sensitivity sample, where we restricted to only those with complete data for active travel behaviour. Compared with the main sample, the sensitivity sample generally showed lower levels of active travel. A greater proportion reported car use, no travel, or not working in the sensitivity sample (94.3% vs. 89.7%). The sensitivity sample also had more participants with journey times of 0 minutes for both outward (81.0% vs. 62.9%) and return journeys (81.1% vs. 62.9%).

**Table S5**

*Distributions across the main analytic sample and the sensitivity sample (with complete active travel data) with regards to both active travel modes and journey times.*

| Variable | Category | Main sample  N (%) | Sensitivity sample N (%) |
| --- | --- | --- | --- |
| **Active travel mode** | Active travel + other modes | 875 (8.5%) | 168 (3.1%) |
|  | Active travel only | 117 (1.1%) | 79 (1.4%) |
|  | Car / none / not working | 9246 (89.7%) | 5187 (94.3%) |
|  | Public transport/other only | 72 (0.7%) | 64 (1.2%) |
| **Journey time (outward)** | 0 minutes | 6485 (62.9%) | 4456 (81.0%) |
|  | 8 minutes | 2029 (19.7%) | 545 (9.9%) |
|  | 22 minutes | 1263 (12.3%) | 335 (6.1%) |
|  | 45 minutes | 473 (4.6%) | 151 (2.7%) |
|  | 60 minutes | 60 (0.6%) | 11 (0.2%) |
| **Journey time (return)** | 0 minutes | 6489 (62.9%) | 4457 (81.1%) |
|  | 8 minutes | 2056 (19.9%) | 549 (10.0%) |
|  | 22 minutes | 1206 (11.7%) | 330 (6.0%) |
|  | 45 minutes | 499 (4.8%) | 150 (2.7%) |
|  | 60 minutes | 60 (0.6%) | 12 (0.2%) |

**Table S6**

*Covariate-unadjusted moderation model examining the association between the built environment at enrolment, active travel at a 2-year follow-up, and depressive symptoms at a 4-year follow-up.*

| Predictor | Standardised Estimate (SE) | *p* value |  |
| --- | --- | --- | --- |
| Active travel | -0.008 (0.01) | .435 |  |
| Built environment | 0.043 (0.01) | < .001 | *** |
| Active travel × built environment | -0.007 (0.01) | .462 |  |
| Adjusted R^2^ | .002 |  |  |

*Note. * p < .05, ** p < .01, *** p < .001.*

**Table S7**

*Covariate-adjusted sensitivity model (complete active travel data only) moderation model examining the association between the built environment (at study enrolment), active travel (at a 2 year -follow-up), and depressive symptoms (at a 4-year follow-up).*

| Predictor | Standardised Estimate (SE) | *p* value |  |
| --- | --- | --- | --- |
| Active travel | 0.002 (0.01) | .856 |  |
| Built environment | 0.017 (0.01) | .151 |  |
| Age at enrolment | 0.050 (0.01) | < .001 | *** |
| Familial depressive history | 0.053 (0.01) | < .001 | *** |
| SES Risk Factors | 0.044 (0.01) | < .001 | *** |
| Physical activity | -0.009 (0.01) | .456 |  |
| Baseline depressive symptoms | 0.422 (0.01) | < .001 | *** |
| Smoking | 0.031 (0.01) | .015 | * |
| Active travel × built environment | -0.009 (0.01) | .519 |  |
| Adjusted R^2^ | .199 |  |  |

*Note. * p < .05, ** p < .01, *** p < .001.*

**Table S8**

*Covariate-unadjusted sensitivity model (complete active travel data only) moderation model examining the association between the built environment(at study enrolment), active travel (at a 2 year -follow-up), and depressive symptoms (at a 4-year follow-up).*

| Predictor | Standardised Estimate (SE) | *p* value |  |
| --- | --- | --- | --- |
| Active travel | -0.004 (0.01) | .787 |  |
| Built environment | 0.036 (0.01) | .007 | ** |
| Active travel × built environment | -0.019 (0.01) | .226 |  |
| Adjusted R^2^ | .001 |  |  |

*Note. * p < .05, ** p < .01, *** p < .001.*

Due to relatively low levels of participants who only engaged with active travel, we additionally ran an exploratory analysis investigating whether we would observe similar findings where active travel behaviour was dichotomised into participants who engaged in active travel and those who did not (**Table S9**). As shown in **Table S10**, participants living in areas characterised by higher built environment factor scores had greater odds of engaging in active travel behaviour (OR = 1.71, 95% CI = 1.55–1.89, *p* < .001).

**Table S9**

*Breakdown of the proportion of participants who engaged in active travel and those who did not.*

| Active travel group | N (Percent) |
| --- | --- |
| No active travel | 9318 (90.4%) |
| Any active travel | 992 (9.6%) |

**Table S10**

*Association between built environment (at study enrolment) and binary active travel (at a 2-year follow-up).*

| Predictor | Odds ratio | 95% CI | *p*-value |  |
| --- | --- | --- | --- | --- |
| Built environment factor | 1.713 | [1.549, 1.894] | < .001 | *** |
| Age at enrolment | 1.012 | [0.998, 1.027] | .088 |  |
| Familial depressive history | 1.148 | [1.004, 1.309] | .041 | * |
| SES risk factors | 1.021 | [0.882, 1.180] | .779 |  |
| Physical activity | 1.008 | [1.004, 1.012] | < .001 | *** |
| Depressive Symptoms at enrolment | 0.982 | [0.960, 1.004] | .111 |  |
| Smoking | 0.965 | [0.881, 1.056] | .437 |  |
| *Note. * p < .05, ** p < .01, *** p < .001.* |  |  |  |  |

**Table S11**

*Covariate-adjusted linear regression examining whether binary active travel behaviour (at a 2 year follow-up) moderates the association between the built environment factor (at study enrolment) and depressive symptoms (at a 4 year follow-up).*

| Predictor | β (SE) | 95% CI | *p*-value |  |
| --- | --- | --- | --- | --- |
| Any active travel (vs. no active travel) | -0.057 (0.03) | [-0.119, -0.005] | .072 |  |
| Built environment factor | 0.024 (0.01) | [0.006, 0.043] | .011 |  |
| Age at enrolment | 0.049 (0.01) | [0.031, 0.067] | <.001 | *** |
| Familial depressive history | 0.115 (0.02) | [0.078, 0.151] | <.001 | *** |
| SES risk factors | 0.044 (0.01) | [0.025, 0.063] | <.001 | *** |
| Physical activity | 0.006 (0.01) | [-0.012, 0.024] | .505 |  |
| Baseline depressive symptoms | 0.399 (0.01) | [0.381, 0.418] | <.001 | *** |
| Smoking | 0.035 (0.01) | [0.011, 0.059] | .004 | ** |
| Any active travel × Built environment factor | -0.001 (0.03) | [-0.06, 0.059] | .996 |  |

*Note. * p < .05, ** p < .01, *** p < .001. Active travel group was a categorical predictor and so was not standardised.*

Exploratory correlation analyses revealed a consistent pattern of results, whereby correlations between six individual built environment indicators (at study enrolment) and active travel behaviour (at 2-year follow-up) were weak (*r* = −0.084 to 0.083, *p* = all < .001), alongside weak correlations between individual built environment indicators (at study enrolment) and depressive symptoms (at 4-year follow-up) (*r* = -0.027 to 0.049, *p* = .449 to < .001) (**Table S12**). Walkability (at study enrolment) showed the strongest correlation with depressive symptoms (at 4-year follow-up) (*r* = 0.049, p < .001). Green space (at study enrolment) showed the strongest correlation with active travel behaviour (at 2-year follow-up) (*r* = -0.084, p < .001).

**Table S12.**

*Exploratory correlation analyses between individual built environment indicators (at study enrolment) with active travel behaviour (at 2-year follow-up) and depressive symptoms (at 4-year follow-up).*

|  | Active Travel Behaviour (2-year follow-up) | | Depressive Symptoms (4-year follow-up) | |
| --- | --- | --- | --- | --- |
|  | *r* | *p* | *r* | *p* |
| Building density | 0.083 | < .001 | 0.017 | .080 |
| Connectivity density | 0.079 | < .001 | 0.034 | < .001 |
| Facility richness | 0.075 | < .001 | 0.036 | < .001 |
| Green space | −0.084 | < .001 | −0.027 | .006 |
| Population density | 0.056 | < .001 | 0.007 | .449 |
| Walkability | 0.074 | < .001 | 0.049 | < .001 |
